# Burden and Predictors of Underweight among Preschool Orphan Children in Southern Ethiopia

**DOI:** 10.1101/2020.10.24.20218768

**Authors:** Adane Tesfaye, Andnet Tadesse Wete, Belay Negassa, Yawkal Chane, Tekle Ejajo, Abebaw Molla, Alemu Basazin Mingude, Lemma Getacher

**Affiliations:** Department of Human Nutrition, School of Public Health, College of Medicine and Health Science, Dilla University, Dilla, Ethiopia; Department of Public Health, College of Medicine and Health Sciences, Dilla University, Dilla, Ethiopia; Department of Nutrition, College of Medicine and Health Science, Kotebe Metropolitan University, Addis Ababa, Ethiopia; Department of Health Service Management, College of Medicine and Health Science, Wachemo University, Hossana, Ethiopia; Department of Public health, College of Health Science, Mizan Tepi University, Mizan Tepi, Ethiopia; Department of Nursing, College of Health Science, Debre Berhan University, Debre Berhan, Ethiopia; Department of Public health, College of Health Science, Debre Berhan University, Debre Berhan, Ethiopia

**Keywords:** Underweight, Preschool, Orphan children, Southern Ethiopia

## Abstract

**Background:** Underweight is one of the public health problems in Ethiopia. Underweight children had lower resistance to diseases, lower school performance, and poor quality of life. In Ethiopia, most of the available evidences are related to the general community children, which had different risks and severity level than orphan children. Even though under-five orphan children had a higher risk of underweight, they are the most neglected population. Therefore, the aim of the study to determine the burden and predictors of underweight among preschool orphan children in Dilla Town, Southern Ethiopia.

**Method:** A community-based cross-sectional study was conducted among 367 orphans from December 5, 2017 to January 30, 2018. The survey data were entered into EPi-info version 3.5.4 software and exported to SPSS version 20 for analysis. The burden of underweight was assessed by calculating the percentages using ENA SMART software was used for anthropometric data management using WHO standard cutoff point below-2 S.D using z-scores. All variables with a p-value of < 0.25 during bivariate logistic regression analysis were entered into a multivariate analysis to identify predictors variables independently associated with underweight at a p-value of 0.05 with 95% CI.

**Results:** In this study, the burden of underweight among orphan children was 27.4%. The main predictors of underweight were sex of child (AOR = 5.29, 95% CI (2.83-9.92)), type of first complementary food (AOR = 2.47; 95% CI (1.24-4.94)), household food security (AOR = 1.98; 95% CI (1.23-3.21)) and age of child (AOR = 7.19; 95% CI (3.81-13.60)).

**Conclusion:** Underweight is a public health problem in the study area. Sex of a child, type of first complimentary food, household food security status, and child age were the predictors of underweight. Therefore, dietary appropriate intervention, nutrition education of mothers, and increase food security status of orphan children are highly recommended.

## BACKGROUND

Underweight is expressed as the percentage of children aged 0 to 5 years whose weight for age is below minus two standard deviations and minus three standard deviations considered as moderate and severe underweight respectively according to the WHO Child Growth Standards. Underweight, based on weight-for-age, is recommended as the indicator to assess changes in the magnitude of malnutrition over time [**1-3**].

Because of the severity of nutritional problems, nutrition is placed at the heart of Sustainable Development Goals (SDG) and vital for achieving 12 out of 17 SDGs, in fact, the remaining 5 SDGs also in one- or another-way support improvements in nutrition. Generally, out of the total 242 indicators of SDGs, 56 are highly relevant to nutrition [**4**].

Worldwide, malnutrition is categorized as the leading cause of morbidity and mortality among under-five children in developing countries which contributes more than 50% of all causes of death. Undernourished children had lowered resistance to diseases, lower school performance, and many other indicators of poor quality of life. Over 90% of all orphans are not living with a surviving parent that believed to suffer more from the effect of undernutrition so far [**5-7**].

Millions of children have been orphaned in the African continent, of these approximately 12.3 million children have been orphaned in sub-Saharan Africa (SSA), because of the impact of AIDS, conflicts, political instability, and other many more reasons. This figure will increase in the next decade as HIV-positive parents become ill and die from AIDS, and vulnerability also keeps increasing because of the above-listed problems [**8, 9**]. Moreover, in SSA, orphan children are more suffering from the impact of undernutrition and more than one-third of countries had a 25% magnitude of underweight [**7**,**10**].

In Ethiopia, around 5.5 million children (6% of the whole population) are categorized as orphans and vulnerable children (OVC), of these 640,802 maternal orphans, 550,300 paternal orphans, and 304,282 dual orphans [**11**]. The burden of underweight among these populations ranges from 8.9% to 27.6% in Ethiopia [**12-15**]. Besides, most of the orphans are given extended family or foster care, and very few are supported by nongovernmental organizations’ care centers found in regional cities. Missing parental care and support increased their vulnerability to food insecurity, chronic undernutrition, acute malnutrition, lack of protection, lack of shelter, lack of education, exposed to physical and sexual abuse in advance [**6, 7**].

According to EDHS 2016, undernutrition is still among the most serious public health problem of under five years children facing in Ethiopia, and among them, orphans are considered the riskiest and vulnerable group of the population. In this survey, 24% of under-five children were underweight [**4**].

The major factors that associated with underweight of orphan children were inadequate dietary intake, disease conditions, absence or limited child care, socio-demographic variables (low income, lack of education, increased family size, age, and marital status), environmental and hygienic conditions (**1, 8, 11, 15, 16**).

In Ethiopia, most of the studies are related to the general community of under-five children, which had different risks, predisposing factors, and severity level than orphan children. Caring for orphan children is one of the intervention strategies of Ethiopia through community-based programs, but it failed to bring the designed impact. Even though under-five orphan children had a higher risk of underweight, they are the most neglected population in Ethiopia. Besides, Dilla Town lacks information concerning the burden of underweight among under five years of orphan children, while this segment of populations is potentially at greater risk of undernutrition due to poor nutrition, less social care, and medical care. Therefore, this study was conducted to determine the burden and potential predictor factors affecting undernutrition among preschool orphan children in Dilla Town, Southern Ethiopia.

## METHODS

### Study Setting and Design

The study was done in Dilla Town, Gedeo zone, Southern Ethiopia. The town is located 359 km from Addis Ababa (the capital city of Ethiopia). The town is bordering with Oromia to the West, Sidama zone to the North, Bule district to the East, and Dilla zuriya district to the south. It is divided into three sub-cities namely, Bedecha, Harowolabu, and Sesa, and also comprises nine kebeles (lowest administrative unit in Ethiopia). Its astronomical location is 6°20’ North Latitude and 38°13’ East Longitude. The total population was 94,189, of which 46,058 (49.9 %) are males and 48,131 (50.1 %) are females. Though the total number of orphans in Dilla town was not known before, according to complete enumeration carried out for the sake of this study, it was 2895. Community-based cross-sectional study design was employed.

### Study Population

All preschool orphan children aged 6-59 months were the source population. While the study population for this study was randomly selected orphan children aged 6-59 months in selected kebeles of Dilla town. Orphan children aged 6-59 months who were severely ill, lived for less than six months and children with immobility precautions like fracture were excluded. The sample size was calculated considering, the proportion of underweight (31.7%) [**3**], 95% confidence interval, and a 5% marginal error, and by adding a 10% non-response rate. Accordingly, the calculated sample size for this study was 367.

### Data Collection Instruments and Procedures

To collect the data for this study, a pre-tested interviewer-administered questionnaire was used. It was further adapted and modified from various relevant studies that meet the aim of the study. First, the questionnaire was prepared in English language and translated into Amharic and Gedeaufa (local languages) to obtain necessary information on demographic characteristics, socioeconomic status, immunization status, food security status, sanitation and hygienic conditions, and feeding practices and child care of preschool orphan children.

The data were collected by six data collectors who were diploma nurses and supervised by two health officers. Data collectors and supervisors were given two days of training by the principal investigator. The data collection, application of standard procedures, and accuracy of results were supervised and checked daily for its completeness and consistency through strong follow up by the principal investigator. For anthropometric measurements, the average result has been considered after repeated measurements. The weight of the child was measured to the nearest 0.1 kg using a 25 kg hanging spring scale with light clothes and without shoes.

### Data Processing and Analysis

Data were entered and cleaned up using EPI-info version 3.5.4 and ENA for SMART 2012 version software was used for anthropometric data management. Logistic regression analysis using SPSS Windows version 20 was done. After having all variables with p-value < 0.25 as a candidate during bivariate analysis; multivariate analysis was carried out to see the effect of each independent variable on nutritional status explained as Underweight at p-value 0.05 with 95% CI. Principal component analysis (PCA) was carried out for the reduction of variables involved in wealth index, food security status, and balanced diversified diet assessment.

### Ethical considerations

The study was ethically approved by the Institution Review Board (IRB) of the College of health science, Dilla University. It was approved on December 2, 2018 and numbered with a Ref C/AC/R/D/987/18. And permission letter for data collection was received from the Dilla town health Bureau. The mothers/caregiver of each study participant were informed about the research objectives, methods, and techniques in detail and informed consent was taken. And then the written and signed consent was obtained from each study participant’s parents. The confidentiality was maintained as appropriate. As a value-adding consideration, nutrition education was given to study participant mothers/caregivers.

## RESULTS

### Socio-demographic characteristics of the respondents

In this study, a total of 361 preschool orphan children were participated making a response rate of 98.4%. Of these, 189 (52.4%) were female by sex, 202 (56%) were maternal orphan children, 160 (44.3%) were in the age category 48-59 months with a mean age of 42.55 months, 292 (80.9%) were in a household who had number of under-five children greater or equal to two, and 140 (38.8%) of orphan children care giver were not attend any formal education (**Table 1**). Besides, the details of the socio-demographic characteristics of participants have been described elsewhere in an earlier study from the same research project [**17**].

**Table 1:** Sociodemographic characteristics of respondents in Southern Ethiopia, 2018 (n= 361)

| **Variables** | **Category** | **Frequency** | **Percent (%)** |
| --- | --- | --- | --- |
| **Sex of a child** | Male | 172 | 47.6 |
|  | Female | 189 | 52.4 |
| **Status of Orphans children** | Double orphans | 67 | 18.6 |
|  | Maternal orphans | 202 | 56.0 |
|  | Paternal orphans | 92 | 25.5 |
| **Age of a child in a month** | 6-11 | 1 | 0.3 |
|  | 12-23 | 17 | 4.7 |
|  | 24-35 | 96 | 26.6 |
|  | 36-47 | 87 | 24.1 |
|  | 48-59 | 160 | 44.3 |
| **Number of Under-five Children in HH\*\*** | <2 | 292 | 80.9 |
|  | ≥2 | 69 | 19.1 |
| **Sex of a caregiver** | Male | 19 | 5.3 |
|  | Female | 342 | 94.7 |
| **Age of a caregiver** | 18-49 | 302 | 83.7 |
|  | ≥50 | 59 | 16.3 |
| **Head of a family** | Mother | 112 | 31.0 |
|  | Father | 233 | 64.5 |
|  | Other *** | 16 | 4.5 |
| **Education status of a caregiver** | No formal education | 140 | 38.8 |
|  | Grade 1-8 | 106 | 29.4 |
|  | Grade 9-12 | 76 | 21.1 |
|  | Above grade 12 | 39 | 10.8 |

\*uncles, aunts, \*\*HH-household, \*\*\*Firstborn, uncles, aunts, in-laws,

### Sanitation and hygiene characteristics

In this study, the source of water for orphan children was public stand 229 (63.4%) and followed by pipe water119 (33.0%). The amount of water used per day >15L was 326 (90.3%) of households. The majority of the households 344 (95.3%) used containers as a means of water storage. Approximately 98.6% of the household had a trend of handwashing with soap while feeding a child. About 93.9% of the household had a latrine facility, of these, pit latrine accounts (49.6%) followed by 36.0% of VIL (**Table 2**).

**Table 2.** Sanitation and hygiene characteristics of the study participants in Southern Ethiopia.

| **Variables** | **Categories** | **Frequency** | **Percent (%)** |
| --- | --- | --- | --- |
| **Water source** | Pipe | 119 | 33.0 |
|  | Public stand | 229 | 63.4 |
|  | Protected spring/well | 13 | 3.6 |
| **Water amount used in a day** | ≤15L | 35 | 9.7 |
|  | >15L | 326 | 90.3 |
| **Water storage method** | Pot | 17 | 4.7 |
|  | Container | 344 | 95.3 |
| **Handwashing with soap while feeding** |  |  |  |
|  | No | 5 | 1.4 |
|  | Yes | 356 | 98.6 |
| **Latrine availability** | Absent | 22 | 6.1 |
|  | Present | 339 | 93.9 |
| **Type of latrine** | Pit latrine | 179 | 49.6 |
|  | VIL | 130 | 36.0 |
|  | Water carriage type | 24 | 6.6 |
|  | Other | 6 | 1.7 |
| **Waste disposal system** | Pit | 113 | 31.3 |
|  | Open space | 21 | 5.8 |
|  | Municipality service | 227 | 62.9 |
| **A separate room used for the kitchen** |  |  |  |
|  | Absent | 317 | 87.8 |
|  | Present | 44 | 12.2 |

### Household Food Security Characteristics

Worrying about not having enough food, not eat preferred food, eat just a few foods, and eat food that you preferred not to eat due to lack of resources were the most frequently occurred variables during household food security status analysis (**Table 3**).

**Table 3:**
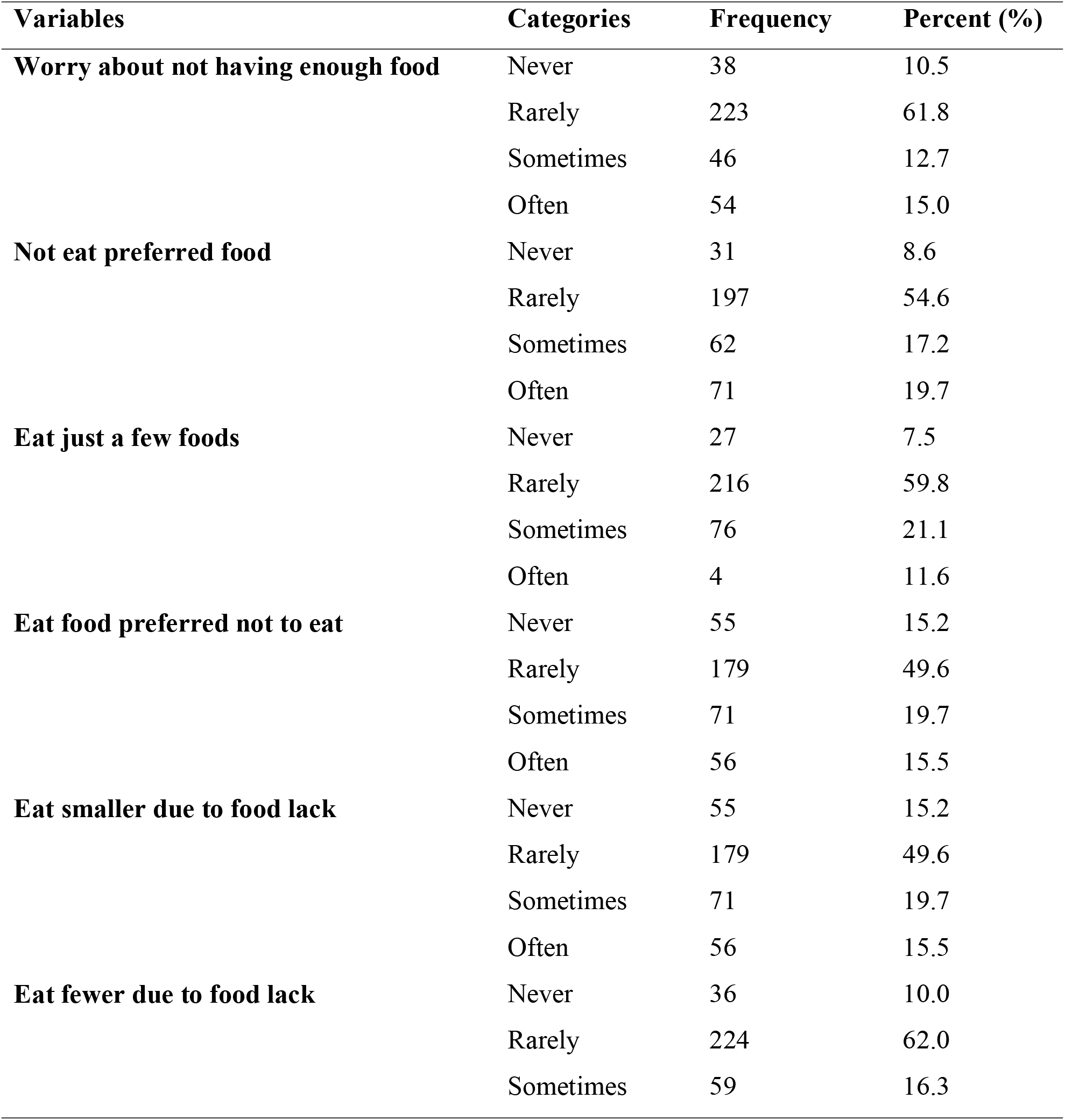
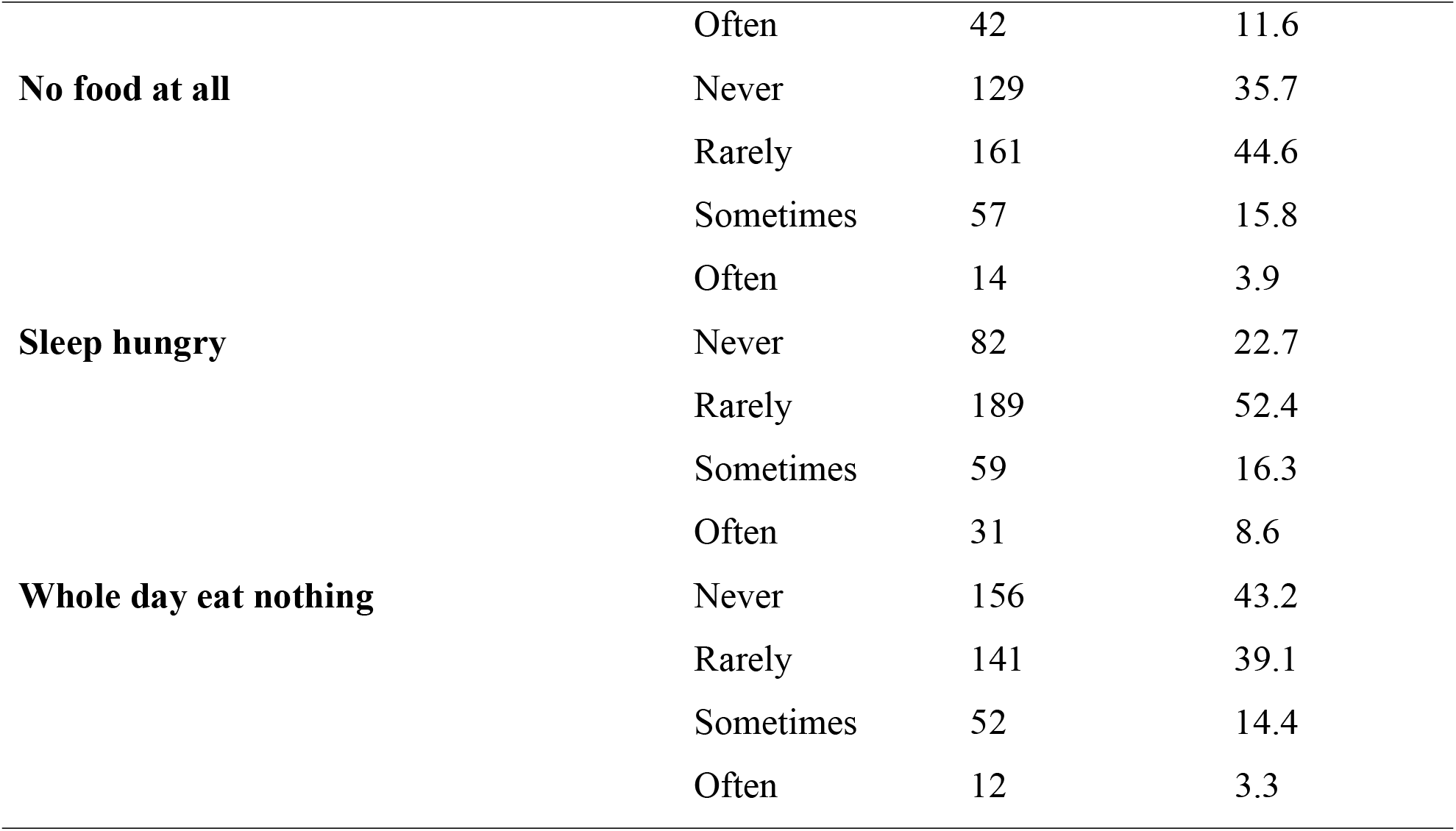
Responses related to household food security characteristics in Southern Ethiopia, 2018.

### Burden and Predictor of Underweight

In the present study, more than quarter (27.4%) of preschool orphan children were affected by the high burden of underweight. In the bivariate logistic regression analysis, those variables with a P-value of ≤ 0.25 were further entered to multivariate logistic regression analysis to identify the independent predictor of underweight. After adjusting for all possible confounders, sex of a child, the type of first complementary food, food-insecure household, and child age were the independent predictor of underweight **(Table 4)**.

**Table 4:** Bivariate and multivariate logistic regression analysis of factors associated with underweight among preschool orphans in southern Ethiopia, 2018.

| Variable | Underweight |  | COR (95% CI) | AOR (95% CI) |
| --- | --- | --- | --- | --- |
|  | Yes (%) | No (%) |  |  |
| Sex of child |  |  |  |  |
| Male | 20 (11.63) | 152 (88.37) | 1 | 1 |
| Female | 80 (42.33) | 109 (57.67) | 5.58 (3.23-9.65) ** | 5.29 (2.83-9.93) ** |
| Alive parent |  |  |  |  |
| No one | 26 (38.81) | 41 (61.19) | 1.542 (1.07-3.21) ** | 1.08 (0.62-1.89) |
| Mom/Dad | 74 (25.17) | 220 (74.83) | 1 | 1 |
| Caregivers' sex |  |  |  |  |
| Male | 8 (42.11) | 11 (57.89) | 1.56 (0.07-2.73) * | 1.61 (0.48-5.45) |
| Female | 92 (26.90) | 250 (73.10) | 1 | 1 |
| Water source |  |  |  |  |
| Pipe | 27 (22.69) | 92 (77.31) | 0.85 (0.55-1.31) * | 0.47 (0.14-1.55) |
| Pub. Stand | 68 (29.69) | 161(70.31) | 0.60 (0.69-1.88) * | 0.67 (0.13-2.14) |
| Pro. Spring | 5 (38.46) | 8 (61.54) | 1 | 1 |
| Hand washing with soap while feeding a child |  |  |  |  |
| Yes | 97 (27.25) | 259(72.75) | 1 | 1 |
| No | 3 (60.00) | 2(40.00) | 2.20 (1.05-4.59) * | 0.47(0.06-3.74) |
| Duration breastfed |  |  |  |  |
| 0-6 month | 56 (31.29) | 123 (68.71) | 1.29 (0.92-1.81) * | 1.71 (0.64-4.54) |
| >6 month | 44 (24.18) | 138 (75.82) | 1 | 1 |
| **Type of first complementary food** |  |  |  |  |
| Milk | 28 (29.79) | 66 (70.21) | 0.76 (0.48-1.21) * | 1.33 (0.77-2.27) |
| Adult food | 18 (40.91) | 26 (59.09) | 1.22 (0.87-1.69) * | **2.47 (1.24-4.94) \*** |
| Porridge | 54 (24.22) | 169 (75.78) | 1 | 1 |
| **Vaccinated** |  |  |  |  |
| Yes | 78 (26.80) | 213 (73.20) | 1 | 1 |
| No | 18 (36.00) | 32 (64.00) | 1.46 (1.65- 7.76) * | 2.25 (0.65-7.76) |
| Not known | 4 (20.00) | 16 (80.00) | 1.34 (0.54-3.28) * | 1.46 (0.46-4.52) |
| **Wealth index** |  |  |  |  |
| Poor | 40 (32.5) | 83 (67.48) | 0.91 (0.80-1.04) * | 1.58 (0.89-2.79) |
| Medium | 32 (27.1) | 86 (72.88) | 0.71 (0.43-1.18) * | 1.22 (0.68-2.19) |
| Rich | 28 (23.3) | 92 (76.67) | 1 | 1 |
| **HH food security status** |  |  |  |  |
| Food secure | 61 (33.70) | 120 (66.30) | 1 | 1 |
| Food insecure | 9 (21.67) | 141 (78.33) | 1.83 (1.14-2.94) * | **1.98 (1.23-3.21) \*** |
| **Child age** |  |  |  |  |
| 0-23 months | 18 (10.11) | 160 (89.89) | 1 | 1 |
| 24-59 months | 82 (44.81) | 101 (55.19) | 7.22 (4.09-12.73) ** | **7.19 (3.80-13.60) \*\*** |
| *P-value < 0.25 in the bivariate analysis ** P-value < = 0.05 in the multivariate analysis |  |  |  |  |

Female preschool orphan children were more than five times more likely to have underweight than male preschool orphan children (AOR = 5.29, 95% CI (2.83-9.92)). The odds of underweight for under five years orphans who consumed adult meal as first complementary food were 2.47 times more likely to have underweight than orphans who consumed porridge as first complimentary food (AOR = 2.4, 95% CI (1.24-4.94)) **(Table 4)**.

Preschool children who had food-insecure households were nearly two times more likely to have underweight compared to children who had food-secure households (AOR = 1.98, 95% CI (1.23-3.21)). Underweight was more than seven times more common among preschool orphan children who were in the category of age range from 24-59 months when compared to their counterparts (AOR =7.2, 95% CI (3.80-13.60)) **(Table 4)**.

## DISCUSSION

In this study, the burden of underweight among preschool orphan children in the study area was 27.7%. The present study also examined the necessary information on socio-demographic characteristics, socio-economic status, immunization status, food security status, sanitation and hygienic conditions, feeding practices, and child care of preschool orphan children. Accordingly, the sex of a child, the type of first complementary food, food-insecure households, and child age were the predictors of underweight among these populations.

This result was a bit higher than the studies done among the orphan and vulnerable children in India (18.2%) [**15**], Malawi (24.6%) [**18**], and Kenya (18.1%) [**19**] and also from the regional prevalence of Ethiopia in Tigray (23%), Oromia (22.5%), SNNPR (21.1%), Gambella (19.4%), Harar (20%) and Addis Ababa (5%) [**16**]. However, the burden of underweight in the present study was found to be a bit lower than studies conducted among similar age groups in South Asia (29.3%) [**20**], Sub-Saharan Africa (28.8%) [**21**], and regional prevalence of Ethiopia in Afar (36.2%), Somali (28.7%) and Benshangul Gumuz (34.3%) [**16**]. This might be due to the difference in study segments, study period, socioeconomic characteristics, health service delivery, and study participant. Another possible variation might be due to the involvement of special segments of the study subject who are on care and support.

The prevalence of underweight was found to be relatively consistent with the regional prevalence of Ethiopia in Amhara (28.4%), Gondar city (27.8%) and Dire Dawa Town (26.2%) respectively [**16, 22**]. This might be due to similarities in socio-economic characteristics and age categories of the study participants.

As to the finding of this result, the odds of underweight for female under five years orphans was more than five folds higher than male under five years orphans. This might be due to gender bias in caregiving. Therefore, this implies that balanced care among male and female orphans is also one of the concerns. Regarding the first complementary feeding, there was a significant association between the first complementary feeding and underweight. The risk of underweight for under five years orphans who consumed adult meals as the first complementary food was nearly three times higher than under five years orphans who consumed porridge as the first complementary food. This might be due to the physiologic incompetence of orphans’ gut to tolerate adult meals and end up with underweight secondary to infection and nutritional quality.

The under-five year’s orphans whose caregivers were in the category of the food-insecure household were almost two times prone to underweight than orphans live in food secured. This could be due to the economic reason that household food insecurity is increased among households’ orphans live. It has also been demonstrated that orphaned children in sub-Saharan Africa tend to have a greater risk of undernutrition compared to non-orphans. A similar study conducted in Kenya revealed that food unsecured households were five times more likely to have underweighted children than food secured households [**19**].

The odds of underweight for under five orphans who were in the category of age range from 24-47 month was more than seven times higher than under five years orphans who were in the category of the age range 6-23 month and 48-59 month. The finding of the study is almost identical to the EDHS, 2016, and a study conducted on nutritional status of under five years in Gondar city referring age group 24-35 months. This might be due to the late initiation of complementary feeding and poor knowledge on how to feed young children [**16, 22**]. This implies that awareness creation on feeding and age-related intervention is one of the concerns.

## CONCLUSION

The study revealed that the prevalence of underweight is high among under-five orphans in Dilla town. Sex of a child, first complementary feeding, household food security status, and child age were factors that were associated with underweight. Therefore, preschool orphan children nutrition issues have to be the central of planning and policy making issue of policy makers to save future generation and great attention has to be paid at level, districts to federal. Moreover, community-based nutrition program targeting preschool orphan children should be established to tackle the problem of undernutrition among these population at the individual and community level at large.

## Data Availability

The datasets used and/or analyzed during the current study are available from the corresponding author on reasonable request.

## Acknowledgments

We would like to express our heartfelt gratitude to study participants; data collectors and supervisors. Moreover, we would like to extend our appreciation for Dilla University College of health sciences and referral Hospitals especially for the public health department, for their effort and ongoing coordination for the effectiveness of this study.

## Ethics approval

The study was approved by the Institution Review Board (IRB) of Dilla University College of health science. It was approved on December 2, 2018 and numbered with a Ref C/AC/R/D/987/18. A mother or father or caregiver of each study participant was informed about the research objectives, methods, and techniques in detail and informed consent was taken. And then the consent was ensured and confidentiality was maintained.

## Patient and public involvement

Patients and/or the public were not involved in the design, or conduct, or reporting, or dissemination plans of this research.

## Patient consent for publication

Not required.

## Provenance and peer review

Not commissioned; externally peer reviewed.

## Contributors

AT is involved in the design, data collection, and manuscript writing. ATW, BR, YC, TE, ABM, AM, and LG involved in data analysis, preparing, and editing of the manuscript. Finally, all authors read and approved the final draft of the manuscript.

## Open access

This is an open access article distributed in accordance with the Creative Commons Attribution Non Commercial (CC BY-NC 4.0) license, which permits others to distribute, remix, adapt, build upon this work non-commercially, and license their derivative works on different terms, provided the original work is properly cited, appropriate credit is given, any changes made indicated, and the use is non-commercial. See: http://creativecommons.org/licenses/by-nc/4.0/

